# Intermittent Theta Burst Stimulation Modulates Overlapping Network Architecture Through Neurochemical and Gene-Regulatory Pathways in Autism Spectrum Disorder With Insomnia

**DOI:** 10.64898/2025.12.03.25341545

**Authors:** Huashuang Zhang, Bincan Xiong, Hongchi Liu, Jingwen Zhang, Ruimin Yu, Wenqi Jiang, Jiongjia She

## Abstract

**Background:** Overlapping system-level architecture in the brain, supporting both functional integration and multifunctional regional engagement, may serve as a candidate biomarker for autism spectrum disorder (ASD) interventions. However, the biological mechanisms underlying the modulation of intermittent theta-burst stimulation (iTBS) for the overlapping system patterns are poorly characterized.

**Methods:** Seventy ASD patients with chronic insomnia were randomly allocated to real or sham iTBS group targeting the left orbitofrontal cortex, with one session per day for 8 weeks. Insomnia Severity Index (ISI) and functional magnetic resonance imaging (fMRI) were collected at baseline and post-intervention. To explore the brain overlapping system, the Shannon-entropy diversity coefficient was calculated. Then we used JuSpace toolbox to quantify spatial correlation linking iTBS-induced reorganization to atlas-based nuclear-imaging derived neurotransmitter maps. Transcriptomic–neuroimaging association analysis were subsequently interrogated with Allen Human Brain Atlas gene expression data.

**Results:** Sleep improvement following real iTBS was associated with restored functional integration across 13 multifunctional regions. Furthermore, these changes aligned with mGluR5 and GABA_A_ receptor densities. The overlapping regions were strongly linked to glutamatergic synaptic transmission and calcium-dependent pathways, enriched in cortical excitatory neurons with peak expression during adolescence, and converged into a highly interconnected protein–protein interaction (PPI) network centered on the Ca²⁺–PKA signaling axis.

**Conclusions:** This study provides multi-scale evidence for the biological mechanisms underlying iTBS modulation of the overlapping system patterns among ASD with chronic insomnia.

## Introduction

Autism spectrum disorder (ASD) remains a major challenge for therapeutic development due to its complex neurobiological underpinnings and heterogeneous clinical presentation. Increasing evidence indicates that chronic insomnia is not merely a comorbid condition but is integral to the core pathophysiology of ASD, contributing to neural dysregulation across multiple systems [1–3]. Addressing these sleep-related abnormalities is therefore essential for improving clinical outcomes and advancing mechanistic understanding of ASD. Intermittent theta-burst stimulation (iTBS), a non-invasive neuromodulatory technique capable of inducing long-term potentiation (LTP)-like synaptic plasticity, offers a promising avenue to restore the excitation-inhibition balance and normalize network communication [4,5]. Understanding the brain network mechanisms mediating iTBS-induced modulation represents a crucial step toward optimizing its therapeutic efficacy for ASD.

Growing neuroimaging evidence conceptualises ASD as a disconnection syndrome involving disrupted interactions among large-scale brain systems [6–8]. However, many earlier connectome investigations based on non-overlapping network models have implicitly treated each brain region as belonging to a single, discrete functional network, thereby failing to account for spatially overlapping network organization [9]. In contrast, evidence from real-world neural architectures demonstrates that brain regions commonly contribute to multiple subnetworks, forming an overlapping topology in which nodes participate in several functional circuits [9]. This overlapping architecture reflects regions that are jointly engaged by different functional systems or cognitive processes across both task states and rest. Such regions typically show cross-network coactivation, hub-like structural properties, and considerable dynamic adaptability. Disruption of this architecture in neuropsychiatric conditions has been associated with hub vulnerability and impaired large-scale network integration [10,11]. In the present study, we developed a novel framework to model the brain’s overlapping system-level architecture using the Shannon-entropy-based diversity coefficient, an information-theoretic metric quantifying the heterogeneity of a node’s inter-network connections [12,13]. A higher diversity coefficient indicates more uniform connections across multiple networks, suggesting a bridging role in cross-network information integration. Conversely, a lower value implies that the node primarily interacts within limited networks, reflecting greater functional specialization. This Shannon-entropy diversity coefficient metric enables the identification of key intermediary nodes that may contribute critically to large-scale communication processes and could help elucidate integrative deficits associated with ASD.

Integrating neuroimaging data with neurotransmitter systems offers a powerful framework for elucidating the biological mechanisms of brain disorders [14–16]. Evidence increasingly shows functional heterogeneity in the human brain is constrained by its intrinsic chemoarchitectural scaffold [17]. An imbalance between excitation and inhibition (E/I) has been recognized as a shared pathophysiological mechanism underlying both ASD and insomnia [18]. This imbalance is shaped by regional variations in glutamatergic (mGluR5) and GABAergic (GABA_A_) receptor densities, which modulate local E/I balance and consequently large-scale network connectivity [19]. Our previous work demonstrated that brain stimulation can alleviate insomnia symptoms by regulating GABAergic neurotransmission [20], underscoring the therapeutic relevance of targeting the E/I system. The JuSpace toolbox, which incorporates normative positron emission tomography (PET)-derived neurotransmitter maps, enables testing whether ASD-related network alterations align with underlying neurotransmitter architectures, thereby facilitating cross-scale analyses of functional-neurochemical correspondence [21]. Building upon this trans-diagnostic framework, we propose that integrating measures of network complexity such as the Shannon-entropy diversity coefficient with neurotransmitter distribution profiles may provide new insights into the neurobiological mechanisms underlying ASD.

Additionally, transcriptome-connectome association studies have established a framework linking macroscale brain networks with microscale transcriptional profiles [22]. This framework has subsequently been applied to a range of neurological and psychiatric conditions, such as major depressive disorder [23] and Parkinson’s disease [24], offering important insights into how gene expression gradients influence disease-related network reorganization. However, no study has yet investigated these transcriptome-connectome associations in ASD with chronic insomnia. The biological mechanisms underlying the alterations in the overlapping system-level architecture remain largely unknown.

Therefore, we conducted a randomized, double-blind, sham-controlled trial involving 70 ASD with chronic insomnia to investigate the neurobiological mechanisms underlying the therapeutic effects of iTBS. Specifically, we aimed to test three hypotheses: (1) iTBS induces reorganization of the brain’s overlapping system-level architecture, which exhibits regional specificity across large-scale functional networks and is associated with improvements in insomnia symptoms in ASD. (2) The spatial pattern of iTBS-induced network reorganization corresponds to regional densities of glutamatergic (mGluR5) and GABAergic (GABA_A_) receptors, reflecting modulation of the E/I balance. (3) The spatial distribution of iTBS-related alterations in network integration is correlated with the receptor topography and expression maps of specific genes, providing molecular insights into the multiscale mechanisms underlying iTBS modulation in ASD with chronic insomnia.

## Methods Study

### Design

This research implemented an 8-week randomized, double-blind, controlled design involving two parallel groups: (A) real iTBS and (B) sham iTBS. For all outcomes, assessments were conducted at baseline and repeated after the completion of the 8-week course of intervention. Specifically, the Insomnia Severity Index (ISI) and functional magnetic resonance imaging (fMRI) were administered at baseline and post-intervention. The stimulation was delivered once daily, five consecutive days per week. The study protocol and all accompanying documentation were approved by the Ethics Committee of the Third People’s Hospital of Songzi. The specific analytic framework is shown in **Fig. 1**.

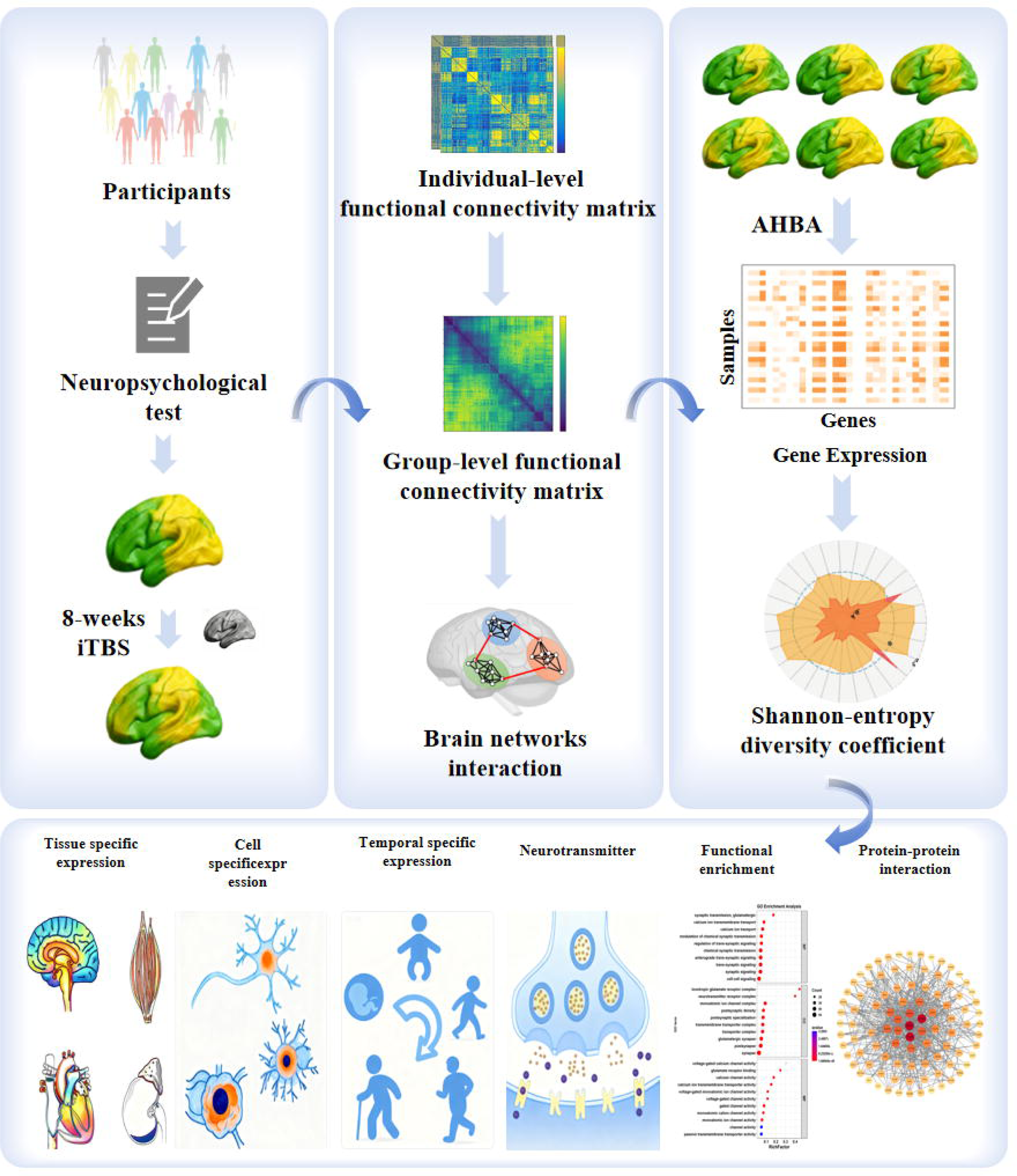

### Participants

The recruitment of participants from the local community was conducted between March 2024 and July 2025, using advertisements, public lectures, and clinical referrals. Eligibility screening was performed by physicians at the TPHS, China. Individuals who met the inclusion criteria were invited to an informational session, during which written informed consent was obtained. Inclusion criteria required: (1) an age range of 18–60 years; (2) a clinical diagnosis of ASD in accordance with DSM-5 criteria [4]; (3) a DSM-5 clinical diagnosis of chronic insomnia; (4) ISI scoring > 7 prior to therapy; (5) Dosage of hypnotics remained stable for the initial 4 treatment weeks; (6) right hand dominance; (7) signed a written informed consent form. The exclusion criteria were: (1) individuals with other untreated sleep disorders including obstructive sleep apnea or hypersomnia.; (2) individuals who are pregnant or lactating; (3) currently or previously participated in other trials; (4) individuals with a history of seizures or serious head injury; (5) intellectual disability (IQ ≤ 70); (6) severe language impairment.

### Sample size

The required sample size was estimated in advance using G*Power 3.1 [25] for a 2 × 2 repeated-measures ANOVA. Assuming a medium effect size according to Cohen’s criteria (f = 0.25, corresponding to ŋ² ≈ 0.06), a significance level of α = 0.05, 90% statistical power, two assessment timepoints, and an expected within-subject correlation of 0.5, the analysis yielded a minimum total sample of 44 individuals (22 per arm). Allowing for an anticipated 20% dropout rate, the target enrollment was increased to 27 participants per group. To further safeguard the study’s power in the event of greater attrition, we ultimately recruited 35 participants for each arm.

### Randomization and Blinding

Eligible participants were assigned to either the real iTBS or sham iTBS group in a 1:1 ratio at random. Randomization was conducted using a computer-generated sequence of random numbers prepared by a statistician blinded to the experimental procedures. Both participants and investigators responsible for assessment were kept blind to treatment allocation. The operator administering iTBS was necessarily unblinded due to the technical requirements of stimulation delivery. To preserve blinding integrity, participants were informed that they would receive iTBS but were not told whether it was active or sham stimulation. They were also instructed not to discuss treatment details with blinded assessors. Blinding efficacy was evaluated after treatment completion by asking participants to guess their group assignment. Accuracy did not exceed chance level (*p* > 0.100), confirming effective masking.

### Intervention: iTBS

Resting motor threshold (RMT) was determined according to established procedures [26]. The figure-of-eight coil (YIRUIDE Company Limited) was positioned stereotaxically over a neuronavigation-defined left orbitofrontal cortex (OFC) anatomical target **(see Supplementary Methods for navigation details).**

The iTBS protocols were adapted from established clinical conventions to enhance participant tolerability and compliance, as demonstrated in prior studies with clinical populations [27, 28]. Each iTBS session commenced with a ramp-up block (600 pulses), during which the stimulation intensity was gradually increased from 0% to the target amplitude, ensuring that only a minimal number of pulses were delivered at a fully efficacious level. The active stimulation block (600 pulses) was then administered at 110% of each participant’s RMT, in accordance with prior protocols [29], or at the highest amplitude tolerated during the ramp-up procedure. Drawing on evidence from TBS clinical trials [30, 31] and electric-field modeling, amplitudes of ≥85% RMT were considered sufficient to elicit meaningful neuromodulatory effects. Participants who were unable to complete the real iTBS block at or above this threshold were excluded from the final dataset.

Across all sessions, stimulation followed a standard theta-burst configuration, consisting of triplet bursts at 50 Hz delivered at a 5 Hz repetition rate. The real iTBS protocol included 20 trains, each 2 s in duration with an 8 s inter-train pause, producing a total stimulation time of 192 s per session. For the sham condition, the identical stimulation protocol was applied over the same target but at 20% of the maximum stimulator output. This corresponded to 32-57% RMT, a level below the threshold for inducing reliable cortical effects, thereby ensuring comparable scalp sensation and auditory cues between active and sham conditions without producing significant neurophysiological changes. The total treatment cycle spanned 8 weeks, with participants receiving one session per day, five days per week, for a total of 40 sessions.

### Outcome measures

#### Primary outcome: fMRI

The Philips 3.0 T MRI scanner (Philips Medical Systems) was used to acquire all fMRI. And the parameters for imaging were as follows: repetition time (TR) = 2000 ms, echo time (TE) = 30 ms, slice thickness = 4.0 mm, flip angle = 90°, matrix size = 64 × 64, number of slices = 35, and spatial resolution = 3 × 3 × 3 mm³.

#### Secondary outcome: ISI

The ISI is a 7-item self-report measure commonly used to assess the severity of insomnia-related symptoms across nighttime difficulties and daytime functioning [32]. Each item is rated on a 5-point Likert scale from 0 (no difficulty) to 4 (very severe difficulty), yielding a total score ranging from 0 to 28. Standard interpretive ranges classify scores of 0-7 as no clinically significant insomnia, 8-14 as mild or subthreshold insomnia, 15-21 as moderate insomnia, and 22-28 as severe insomnia [33]. It is an effective and dependable intervention, notably characterized by its susceptibility to variations in insomnia [34, 35].

### Neuroimaging data analysis

#### fMRI preprocessing

Resting-state data were preprocessed using the GRETNA toolbox [36] running within the SPM12 environment. To reduce magnetization instability at the beginning of each scan, the initial volumes (first five) were discarded. Head movement was subsequently corrected by realigning all images with a rigid-body algorithm. Participants showing substantial motion—operationalized as either translation >3 mm or mean framewise displacement (FD) above 0.5 mm—were excluded, resulting in the removal of two individuals from the real iTBS arm and one from the sham arm. After realignment, the functional series underwent temporal filtering (0.01–0.08 Hz) and nuisance regression using a unified linear model. This model included the 24-parameter motion expansion, along with time courses extracted from white-matter and cerebrospinal-fluid masks and the whole-brain signal, following recommended procedures to reduce physiological and scanner-related noise [37]. White-matter and cerebrospinal fluid (CSF) masks were derived from SPM12 probabilistic tissue priors (threshold = 0.9), and the global signal was taken from the default whole-brain template. Finally, functional images were spatially normalized to the Montreal Neurological Institute (MNI) template by applying the deformation fields estimated during each participant’s structural segmentation, and the normalized images were resampled to 3-mm isotropic voxels.

Shannon-entropy diversity coefficient analysis. Following preprocessing, a functional connectivity (FC) representation was generated for each participant. Cortical time series were extracted based on the 200-parcel Schaefer2018 parcellation, which organizes regions into 17 large-scale functional systems. For each parcel, the mean BOLD signal was obtained, and connectivity estimates were derived by correlating the time courses of all parcel pairs using Pearson correlation. The resulting correlation values were subsequently transformed into z-scores via Fisher’s r-to-z conversion to stabilize variance and approximate normality.

To identify a robust modular structure for subsequent diversity analysis, we employed the Louvain algorithm (resolution parameter γ = 1) from the MATLAB-based brain connectivity toolbox. This algorithm was applied 1000 times to each participant’s FC matrix to account for the stochasticity inherent in the optimization process. To derive a stable consensus partition from these 1000 solutions, a co-assignment matrix was generated, which recorded the frequency with which each pair of nodes was assigned to the same community across all iterations. A final, representative partition for each individual was then obtained by applying a consensus threshold of 0.5 to this co-assignment matrix. Subsequently, group-level consensus partitions were generated by aggregating the individual-level partitions within the real iTBS group and sham iTBS group, respectively.

Finally, based on these stable group-level partitions, the Shannon-entropy diversity coefficient (H) was computed for each node. This coefficient, derived from information theory, quantifies the diversity of a node’s connections across all detected communities, reflecting the extent to which a node participates in multiple overlapping functional systems. The formula for H is as follows:

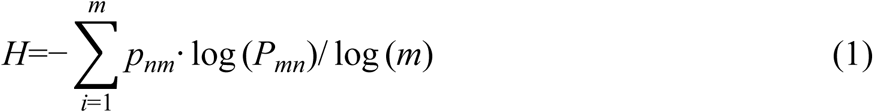

where *P_nm_* represents the proportion of connection strength from node n to module m, and m is the total number of modules.

Identification of differential overlapping regions and links to clinical symptoms. To determine which cortical territories exhibited iTBS-related alterations in system-level integration, diversity coefficient (H) values were contrasted across the real-stimulation and sham conditions (pre- vs. post-treatment). Brain regions surviving a statistical threshold of *p* < 0.05 FDR-corrected) were defined as differential brain overlapping regions.

To investigate whether these iTBS-induced neural changes were linked to clinical improvement, we performed correlation analysis between the changes in regional diversity coefficients (ΔH) and changes in insomnia (ΔISI). The change in diversity coefficient for each significant region was calculated as:

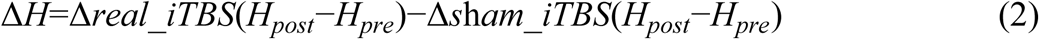

This formula isolates the treatment-specific change by subtracting the mean change observed in the sham group. The change in clinical symptoms was calculated for each participant as:

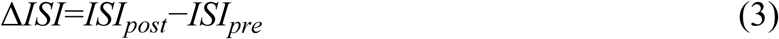

where lower post-treatment scores indicate improvement.

Finally, we computed partial correlations between ΔH and ΔISI across participants, controlling for age, sex, mean FD and baseline ISI scores. The statistical significance of these correlations was assessed using a non-parametric approach with 10,000 permutation tests to obtain robust empirical p values.

### Baseline Overlap-Based Brain Signatures Predict iTBS Efficacy

To determine whether the baseline brain network architecture could predict individual responses to iTBS treatment, we employed a multivariate pattern analysis (MVPA) framework using a linear support vector regression (SVR) algorithm (implemented in LIBSVM [38]). Following previous studies, brain regions that exhibited significant differences in the Shannon-entropy diversity coefficient (H) between the **real** and sham iTBS groups after treatment were defined as masks of interest. We hypothesized that the baseline diversity coefficients within these regions could serve as neural predictors of iTBS therapeutic efficacy. In all participants, the ΔISI was used as the outcome variable, whereas the baseline diversity coefficients (H values) within the identified differential regions were used as predictive features. To mitigate the influence of potential confounding factors, feature values were residualized against age, sex, mean FD, and baseline ISI scores before model training. The predictive model was trained and evaluated using a leave-one-out cross-validation (LOOCV) framework. In each iteration, data from N–1 participants were used for training and the remaining one participant was used for testing. Within each training fold, features showing a significant correlation (Spearman’s *p* < 0.05, uncorrected) with ΔISI in the training set were retained for model construction to prevent overfitting and information leakage. SVR hyperparameters (cost C and epsilon ε) were optimized using a grid search within the training set. The prediction accuracy of the model was quantified by computing the Pearson’s correlation coefficient (r) between the predicted and actual ΔISI values across all participants. To evaluate the statistical significance of this correlation, a permutation test (5000 iterations) was performed by randomly shuffling the ΔISI labels to generate a null distribution of r values. An observed *p* value < 0.05 (two-tailed) was considered statistically significant.

### Spatial concordance with neurotransmitter receptor maps

We assessed the spatial correspondence between iTBS-related functional reorganization in ASD patients with insomnia and the spatial distributions of six neurotransmitter receptor or transporter systems using JuSpace (v1.5) [21]. Multiple comparisons were controlled using FDR adjustment. Density maps for these molecular systems, including D2R [39], DAT [40], mGluR5 [17], VAChT [17], 5-HT1A [41], and GABA_A_ [19], were derived from positron-emission tomography imaging.

### Transcription–neuroimaging association analysis

#### Gene expression dataset and preprocessing

Gene expression data were obtained from the Allen Human Brain Atlas (AHBA), which comprises over 20,000 genes assayed across 3,702 tissue samples from six postmortem donors. Because right-hemisphere sampling in the AHBA dataset is incomplete (only 4 of 6 donors provided right-hemisphere data), we restricted all transcriptomic analyses to the left hemisphere, following established protocols [42,43]. Accordingly, the regional expression matrix included 100 left-hemisphere cortical parcels from the Schaefer-200 atlas. Preprocessing was conducted using the abagen toolbox [42]. The pipeline included: probe filtering (intensity > background in ≥50% of samples), selection of the most differentially stable probe per gene, registration of samples to the Schaefer-200 atlas (2-mm distance threshold), and donor-wise z-score normalization to control for inter-individual differences.

#### Association between gene expression and differential overlapping regions

To determine which transcripts were linked to iTBS-related alterations in overlapping architecture, we examined the correspondence between regional gene expression profiles and the brain areas showing differential overlap changes between the real and sham stimulation conditions. Specifically, gene expression profiles for the identified differential regions were obtained from the AHBA. We then conducted cross-regional Pearson correlations to examine the association between these expression values and the treatment-related changes in Shannon-entropy diversity coefficients across groups. To address multiple comparisons and mitigate the influence of spatial autocorrelation, a two-step significance testing framework was employed. First, for each gene, we computed a spatial autocorrelation-corrected p-value using a nonparametric spatial permutation test (10,000 rotations of the differential brain map via the spin_test function from the neuromaps toolbox). Rotations were hemisphere-preserving on spherical surfaces; parcels overlapping the medial wall were handled by excluding them from the rotation procedure to maintain the empirical distance-dependence. Second, we applied the FDR correction to these corrected p-values (*p* < 0.05). This combined approach ensures that reported associations are robust against both multiple testing and spatial non-independence.

#### Disease-related differential expression analysis

To test the disorder specificity of the imaging-derived gene associations, we extracted log₂ fold-change (log₂FC) values from the cross-disorder transcriptomic dataset of Gandal et al. [44] covering ASD, major depressive disorder (MDD), schizophrenia (SCZ), bipolar disorder (BD), alcoholism and inflammatory bowel disease (IBD). For each disorder, Spearman’s correlations were computed between the imaging–gene association coefficients of the identified genes and their corresponding log₂FC values. Statistical significance was determined by 10,000 permutation tests with FDR correction (*p* < 0.05). A significant or oppositely directed correlation unique to a specific disorder was interpreted as evidence of disease-specific transcriptional coupling.

#### Gene enrichment analysis

Genes implicated in the preceding association analyses were subjected to functional enrichment procedures. Functional annotation was first conducted using the ToppGene platform [45], an integrated toolset commonly employed for gene-function characterization and candidate-gene prioritization through annotation- and network-based approaches [46]. To characterize the biological roles of these genes, Gene Ontology (GO) enrichment was performed across the domains of biological process, cellular component and molecular function. Tissue-, cell type-, and developmental stage–specific expression patterns were then evaluated using the online Tissue Specific Expression Analysis (TSEA) resource [47] and the Cell Type-Specific Expression Analysis (CTEA) tool [48,49]. These analyses allowed us to determine the iTBS-associated gene expression profiles within distinct tissues, cortical cell classes and developmental periods. Specificity index probabilities (P^SI^ = 0.05, 0.01 and 0.001) were applied to quantify the likelihood that a gene showed selective expression within a given class [50].

#### Protein–protein interaction (PPI) analysis

PPI analyses were conducted using the STRING v12.0 [51] to investigate potential interactions among the associated genes. A PPI network was generated, and hub genes were defined as those falling within the top 30% of node-degree rankings, with a minimum confidence score of 0.45. Furthermore, developmental expression patterns of the identified hub genes were assessed using data from the Human Brain Transcriptome Database [52].

## Results

### Participant characteristics

**Table 1** presents the demographic characteristics of the participants (real iTBS and sham iTBS) at baseline. No significant differences existed between these 2 groups at baseline.

### Clinical outcome

Post-treatment ISI scores were markedly lower in the real iTBS group relative to the sham condition (*p* = 0.007). A significant group × time interaction was observed for ISI improvement (F(1,65) = 12.992, *p* = 0.001). Within-group comparisons revealed a mean decrease of −6.94 in the real-stimulation group (95% CI: −8.16 to −5.72; *p* < 0.001), whereas the sham group exhibited a smaller reduction of −3.54 (95% CI: −5.03 to −2.06; *p* < 0.001).

### Network organization differences before and after iTBS

As a preliminary step prior to analysing the Shannon-entropy diversity coefficient, we reconstructed 17 large-scale cortical networks, including the control (Cont A, B, C), default mode (Default A, B, C), dorsal attention (DorsAttn A, B), limbic (Limbic A, B), salience/ventral attention (SalVentAttn A, B), somatomotor (SomMot A, B), temporal–parietal (TempPar), visual central (VisCent), and visual peripheral (VisPeri) networks. To facilitate visualization, each network within the Schaefer-200 cortical parcellation was color-coded according to its spatial correspondence with the canonical 17-network atlas. The network configurations in the real iTBS group before and after treatment were illustrated in **Fig. 2**. Relative to the sham group, participants receiving real iTBS exhibited marked alterations in functional connectivity (i.e., edges) spanning multiple cortical regions (i.e., nodes). In contrast, the sham iTBS group showed no appreciable differences in network configuration between the pre- and post-treatment sessions.

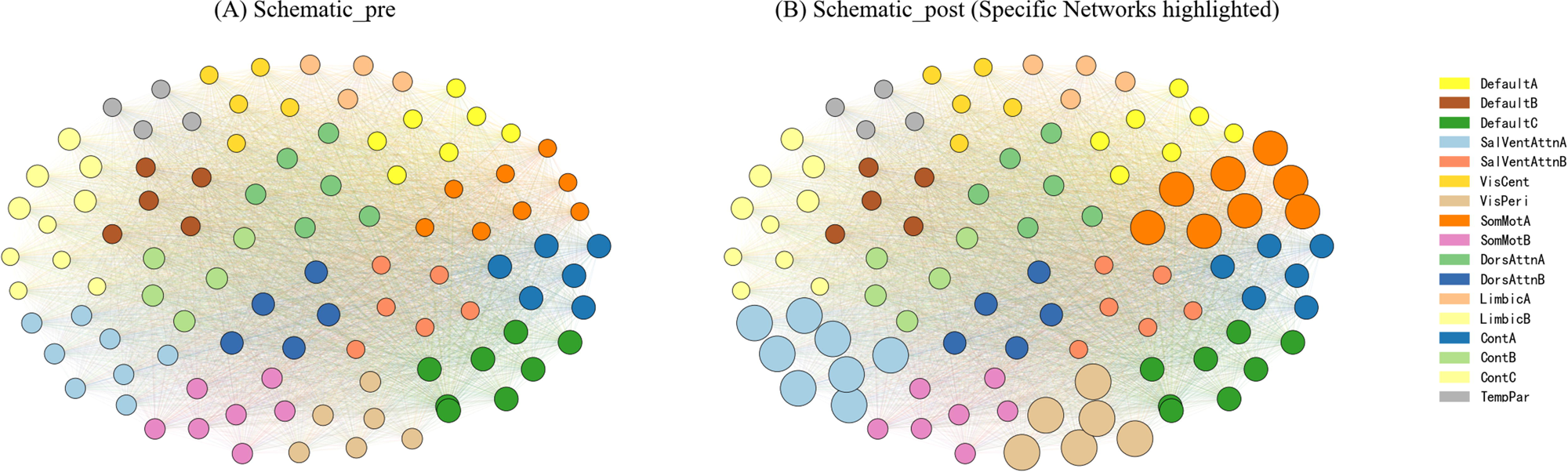

### Differences in overlapping brain architecture between groups

To map alterations in overlapping system organization across the 17 large-scale cortical networks, Shannon-entropy diversity coefficients were computed for every cortical region at both pre- and post-treatment assessments in the real and sham iTBS groups. Group comparisons revealed 13 cortical regions that showed significant treatment-related changes in the real iTBS group relative to the sham group (*p* < 0.05, FDR corrected). These regions were predominantly distributed within the dorsal attention/salience–ventral attention network (DorsAttn/SalVentAttn; n = 5), the somatomotor network (SomMot; n = 4), and the visual network (n = 4) (**Fig. 3**).

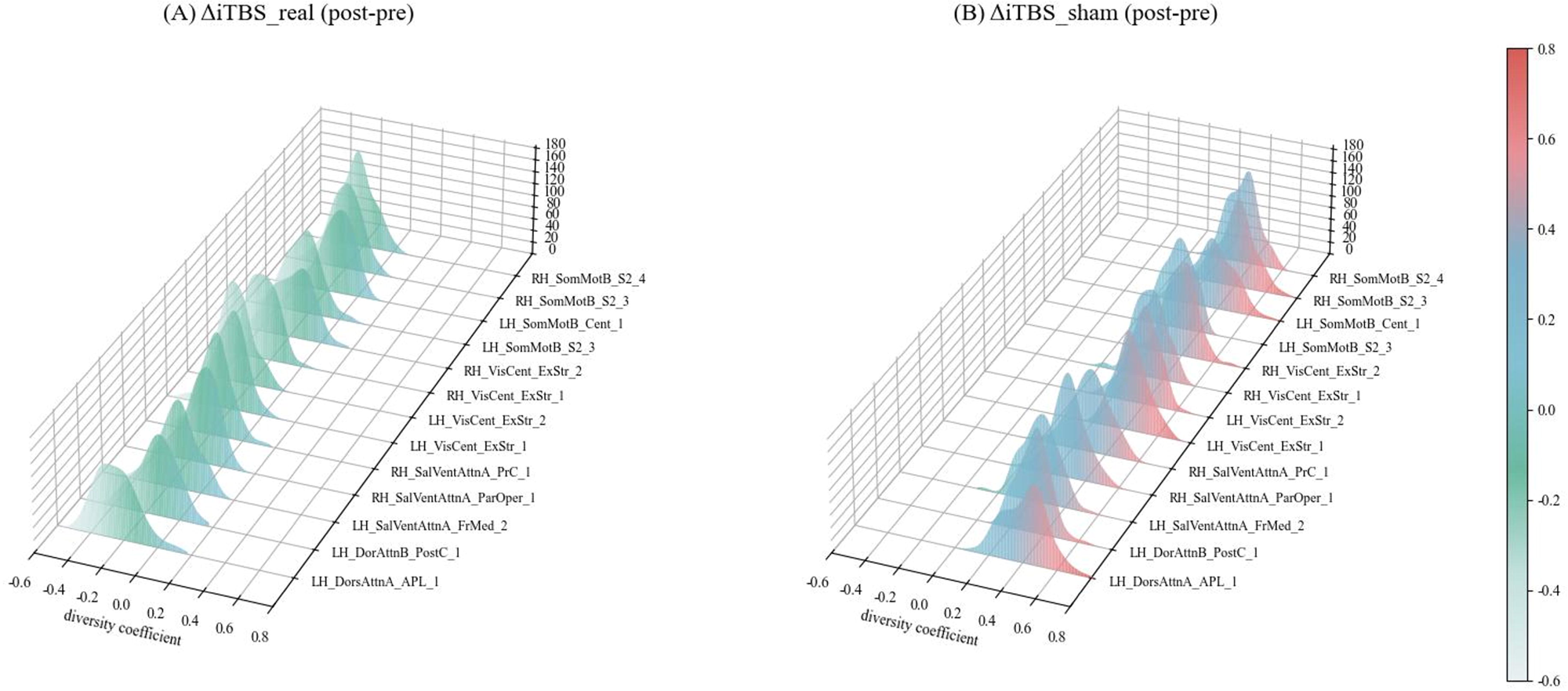

We next examined whether shifts in Shannon-entropy diversity were associated with clinical improvement. After controlling for age, sex, and mean FD, partial correlation analyses indicated that larger iTBS-induced reductions in the diversity coefficient were linked to greater decreases in ISI scores (r = 0.48, *p* = 0.009, FDR corrected). No significant relationships were observed in the sham group.

Given the well-established involvement of the visual network in insomnia pathophysiology [53]; our results further demonstrate that iTBS alters entropy-based diversity within visual regions, and this modulation is positively associated with symptom improvement. Brain regions within the visual circuit that showed a significant real-versus-sham interaction effect (left and right VisCent_ExStr_1; left and right VisCent_ExStr_2) were retained as a mask for subsequent prediction of iTBS treatment responsiveness.

### Predicting iTBS treatment effects from the baseline Shannon-entropy diversity coefficient

At baseline in the real iTBS group, the Shannon-entropy diversity coefficients of the visual-pathway mask regions (left/right VisCent_ExStr_1 and VisCent_ExStr_2) were extracted and used as feature variables. The pre-to-post change in ISI scores was defined as the label. To examine whether these baseline features could predict individual responsiveness to iTBS, we constructed an SVR model incorporating feature-weight–based selection, leave-one-out cross-validation, and permutation testing. Additional experimental results are included in supplementary material (**Fig S1**), baseline entropy coefficients within these regions demonstrated a clear ability to estimate ISI improvement after the 8-week intervention, yielding a correlation of 0.52. Permutation analysis further confirmed that this predictive performance was statistically reliable rather than driven by chance (*p* = 0.001).

### Correlation between Shannon-entropy diversity coefficients and neurotransmitter profiles

The Shannon-entropy diversity coefficients showed significant correlations with two neurotransmitter receptor profiles: mGluR5 (r = 0.215, *p* = 0.002, FDR-corrected) (**Fig. 4C**) and GABA_A_ (r = –0.182, *p* = 0.014, FDR-corrected) (**Fig. 4F**). No significant associations were observed for the remaining neurotransmitter systems, including D2R (r = 0.093, *p* = 0.21) (**Fig. 4A**), DAT (r = 0.086, *p* = 0.24) (**Fig. 4B**), VAChT (r = 0.074, *p* = 0.27) (**Fig. 4D**) and 5HT1a (r = 0.065, *p* = 0.33) (**Fig. 4E**).

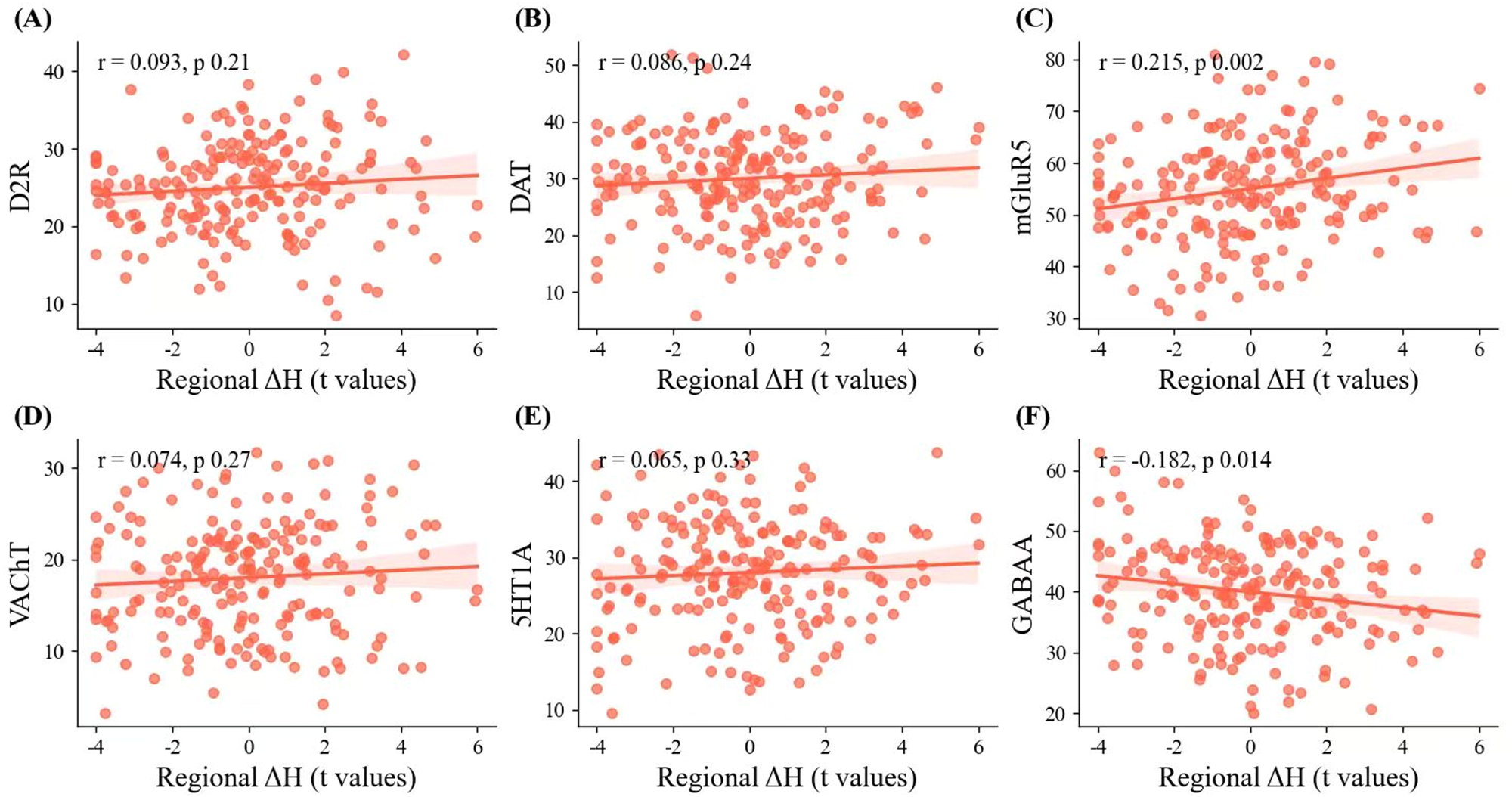

### Association between differential overlapping brain architecture and gene expression

For gene identification, normalized transcriptional data from more than 15,000 genes were obtained from 3,121 tissue samples in the AHBA dataset following relabeling and refined probe selection. These data were then mapped onto cortical parcels to generate a left-hemisphere regional expression matrix. This matrix was used to compute spatial correlations between gene expression and Shannon-entropy diversity coefficients. In the real iTBS group, spatial correlation analyses identified 64 genes (see Supporting Information, Table S1) whose expression patterns significantly matched the spatial distribution of iTBS-related changes in the overlapping brain system (*p* < 0.05, FDR corrected).

### Disease-related differential expression analysis results

We identified 34 genes that were shared between our gene set and those reported by Gandal et al. [44] as significantly dysregulated in postmortem ASD case-control brain tissue. Moreover, the spatial association coefficients of these genes showed a significant negative correlation with their ASD-related expression alterations (r_s(32)_ = −0.39, adjusted *p*_perm_ = 0.012, FDR-corrected) This association was specific to ASD, as it was not observed in the other five disorders: MDD (r_s(152)_ =0.25, adjusted *p*_perm_ = 0.005, FDR-corrected), SCZ (r_s(167)_ = 0.24, adjusted *p*_perm_ = 0.003, FDR-corrected), BD (r_s(96)_ = 0.33, adjusted *p*_perm_ = 0.001, FDR-corrected), alcoholism (r_s(166)_ = 0.06, adjusted *p*_perm_ = 0.150, FDR-corrected), and IBD (r_s(374)_ = 0.07, adjusted *p*_perm_ = 0.180, FDR-corrected) (**Fig. 5**).

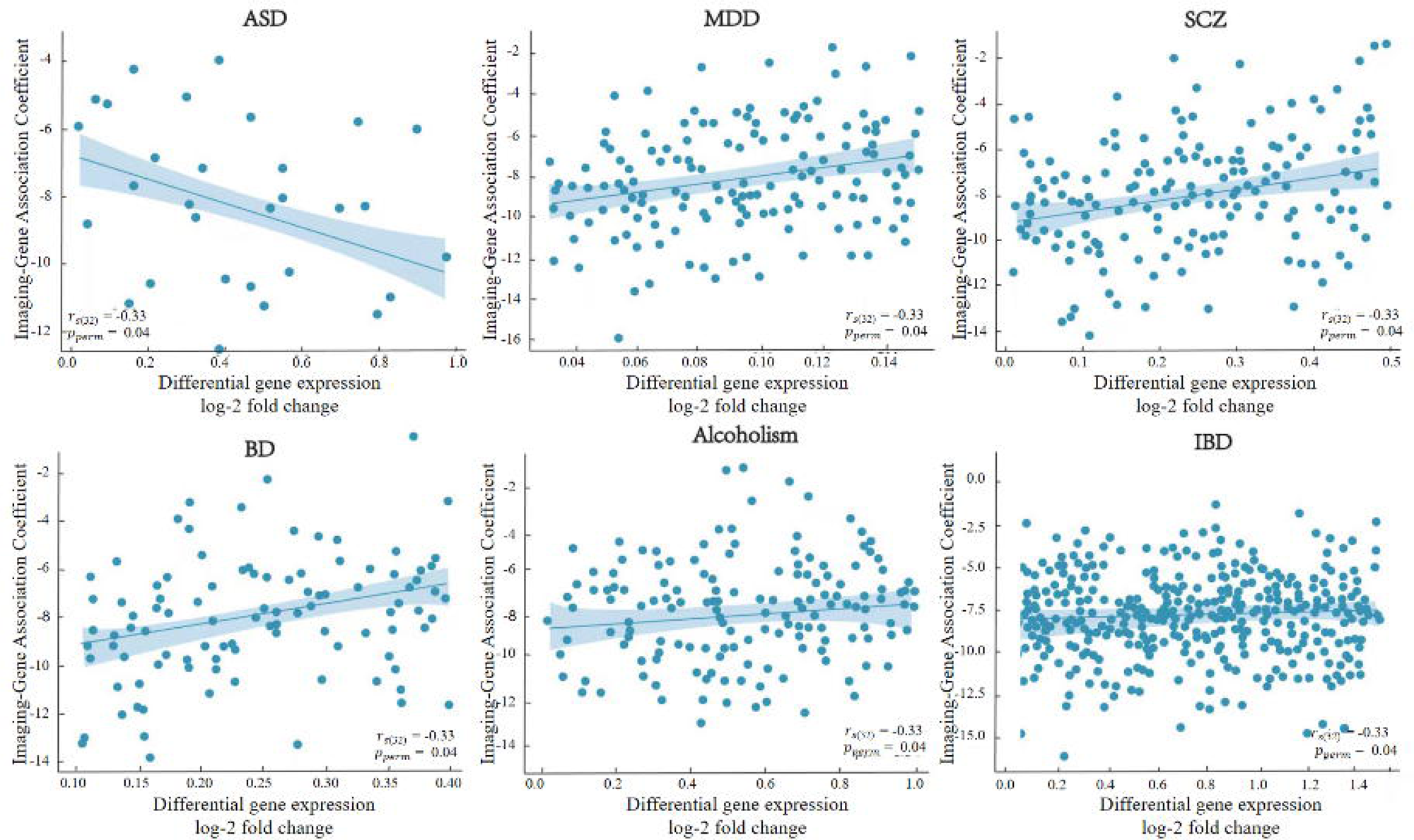

### Gene functional enrichment

To explore the biological significance of the iTBS-sensitive gene set, we performed a comprehensive Gene Ontology (GO) enrichment analysis (**Fig. 6A**). The analysis revealed that the 64 identified genes were significantly enriched in key biological processes such as glutamatergic synaptic transmission and calcium ion transport; cellular components including the postsynaptic density and glutamatergic synapse; and molecular functions such as voltage-gated calcium channel activity and glutamate receptor binding (FDR corrected, *p* < 0.05). These enrichment profiles highlight the strong involvement of synaptic and calcium-related pathways within the iTBS-sensitive gene set.

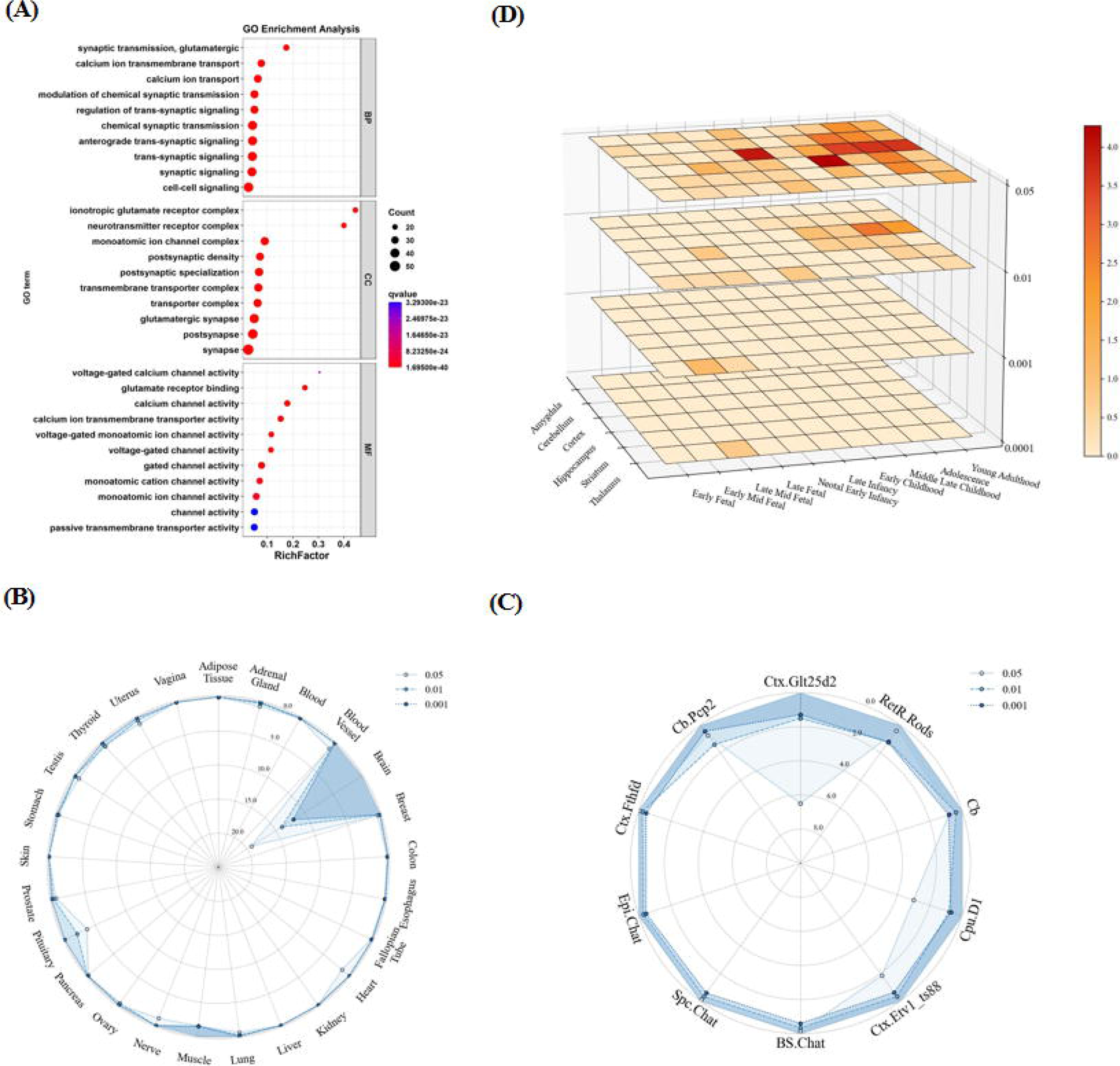

### Tissue, cell type-and temporal-specific expression

Tissue specificity analyses identified the brain as the primary site of expression for these genes. With the highest enrichment observed in cortical and cerebellar regions, followed by hippocampus and striatum (**Fig. 6B**). Cell type–specific profiling further demonstrated significant enrichment within neuronal populations, particularly cortical excitatory neurons (Ctx.Glt25d2) and cerebellar Purkinje cells (Cb.Pcp2), as well as in cholinergic systems across multiple brain regions (**Fig. 6C**). Temporal expression analysis revealed a distinct developmental pattern, characterized by elevated expression during mid-to-late childhood and adolescence, with secondary peaks in early infancy and young adulthood (**Fig.6D**).

### PPI networks and hub genes

PPI analysis revealed a highly interconnected protein–protein interaction network (**Fig.7A**). The resulting network contained 765 edges, significantly greater than the expected 60 edges (PPI enrichment p-value < 1.0 × 10⁻¹⁶), indicating strong functional interconnectivity among these genes. The network exhibited a densely connected core module centered on PRKACB, PRKACA, and PRKACG, which form the catalytic subunits of the cAMP-dependent protein kinase A (PKA) complex. These hub genes were closely linked to calcium channel genes (CACNA1C, CALM1, CAMK2A) and postsynaptic scaffold components (GRIN2B, DLG4). Spatiotemporal expression profiling further demonstrated that all three PRKA genes displayed stable cortical expression across lifespan (**Fig.7B-D**), with pronounced peaks during late childhood and adolescence.

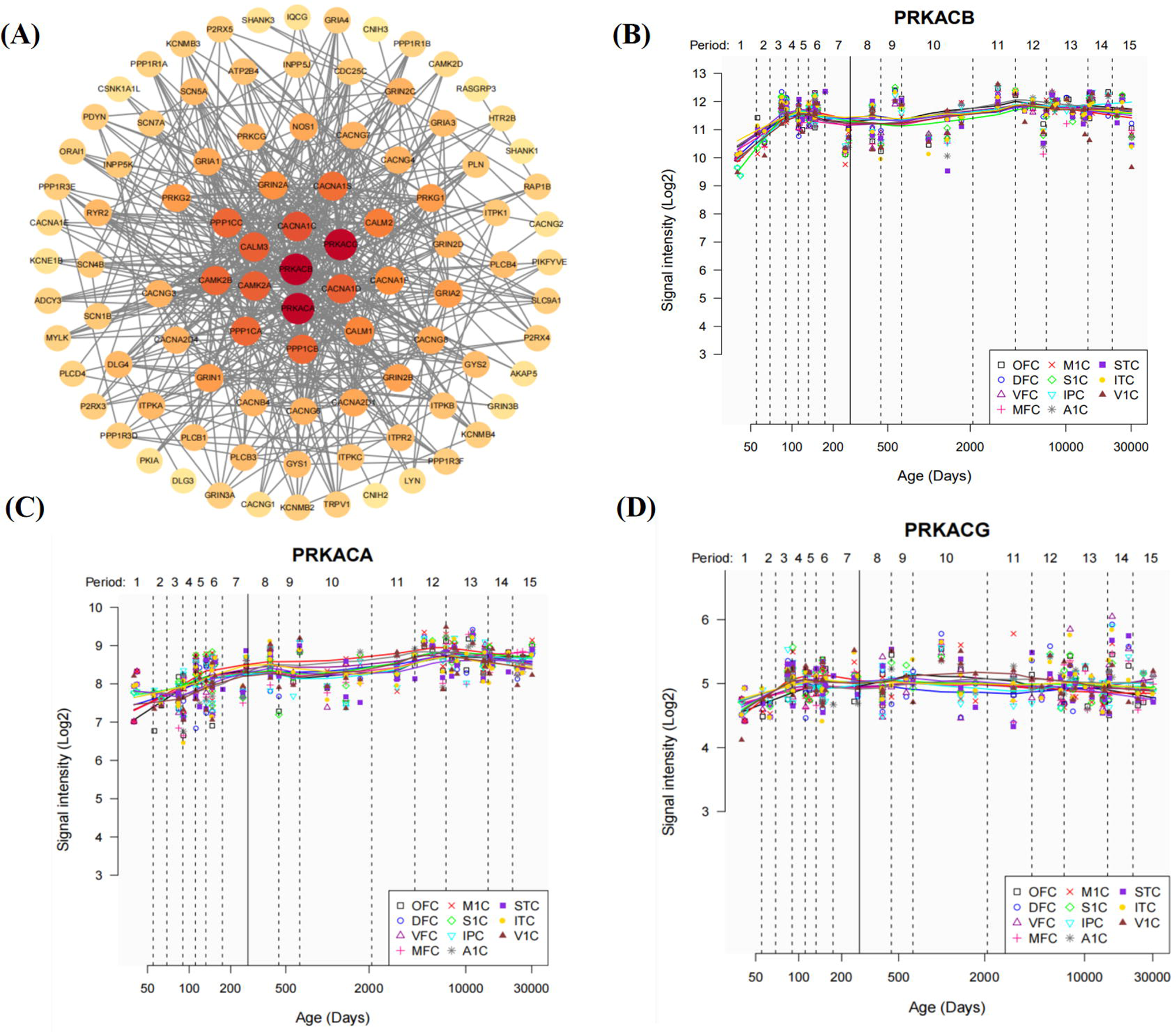

## Discussion

This study demonstrated that an eight-week course of neuronavigated iTBS over the left OFC induced spatially reorganization of the overlapping system-level architecture in ASD with insomnia. The most prominent alterations occurred in the attention, somatomotor, and visual networks and were associated with improvements in insomnia severity. JuSpace analysis revealed that these changes corresponded to regional mGluR5 and GABA_A_ receptor densities, implicating E/I related chemoarchitecture. Transcriptomic profiles further identified convergent gene-expression patterns across iTBS-sensitive regions. To our knowledge, this is the first study to integrate neuroimaging, neurotransmitter mapping and transcriptomic analyses within the framework of the overlapping system-level architecture, providing a multiscale perspective on the biological mechanisms underlying iTBS modulation in ASD with insomnia.

Our findings demonstrate that iTBS induces widespread reorganization of brain networks in ASD with insomnia. This overlapping phenomenon is best characterized as a remodeling of the brain’s overlapping system-level architecture. This observation raises a fundamental challenge to the long-dominant non-overlapping network model which conceptualizes the brain as a set of discrete, unifunctional modules [54,55]. This classical model is hard-pressed to explain how focal stimulation elicits global changes [56]. Our findings strongly align with the emerging conceptual model of widespread overlapping communities in brain networks [9,57], which posits that network overlapping is a fundamental organizational principle, not a temporal byproduct. Within this framework, the system-wide reorganization we observed finds a natural mechanistic explanation: stimulation at a single node may reverberate through the multiple overlapping communities to which it belongs [58]. These observations support the view that cortical regions participate in multiple large-scale systems rather than belonging exclusively to a single network, and that regions with extensive overlap play a key role in facilitating communication across networks [59]. This interpretation is also consistent with evidence that functional networks exhibit flexible and evolving roles, with a splitting phenomenon detected in some of these networks [60]. Consequently, examining the overlapping system-level architecture in ASD with insomnia may advance our understanding of the neurobiological alterations captured by neuroimaging and may offer a potential marker for earlier detection of ASD.

Notably, our spatial mapping analysis revealed a pronounced spatial concordance between the brain regions exhibiting iTBS-induced functional reorganization and the regional densities of glutamatergic [61] and GABA_A_ receptors [19]. This observation points to a potential influence of iTBS are not the result of non-specific local perturbations but rather reflect a targeted modulation of brain networks organized according to specific neurochemical architectures. This interpretation aligns with recent receptor topography research that the spatial distribution of neurotransmitter systems provides a fundamental scaffold for macroscale brain network organization and function [17]. Importantly, this observation is neurobiologically grounded: iTBS has been shown to shift the E/I balance by modulating glutamatergic and GABAergic neurotransmission [62,63] and to induce LTP-like synaptic plasticity through the activation of glutamatergic receptors, including NMDARs and AMPARs [64]. Taken together, these findings indicate that the propagation of iTBS effects across overlapping network topologies is not random but is instead spatially constrained by the brain’s intrinsic chemoarchitectural organization.

Our transcriptomic analysis further bridges these macro- and mesoscopic findings to a core molecular program. The same iTBS-sensitive, receptor-defined regions were enriched for a co-expressed gene set that we term the ‘synaptic tuning toolbox’ functionally associated with synaptic signaling and voltage-gated calcium channel activity. The core protein–protein interaction network of this gene set converged on a Ca²⁺–PKA signaling axis (CACNA1C–CALM1–PRKACB/G), which plausibly mediates the neurophysiological effects of iTBS. Mechanistically, iTBS-induced depolarization and Ca²⁺ influx [65] could be sensed by CALM1, activating downstream PKA catalytic subunits (PRKACB/G) to fine-tune synaptic efficacy—a process consistent with LTP-like plasticity [66]. This integrative framework links the initial electrophysiological events of iTBS to sustained molecular mechanisms of synaptic tuning, further supported by prior evidence of iTBS activating the Ca²⁺–CaMKII–pCREB–BDNF cascade [67,68].

The exceptional efficiency of this molecular toolbox may be rooted in neurodevelopment. Our analysis revealed that its constituent genes exhibit peak expression during late childhood and adolescence—a critical window for experience-dependent synaptic pruning and circuit refinement. This developmental trajectory aligns with large-scale transcriptomic evidence [69,70] and structural co-variation analyses [71], suggesting that this gene program represents an endogenous regulatory mechanism emerging during a phase of heightened plasticity. Previous research has demonstrated that brain stimulation can induce epigenetic modifications influencing gene expression and behavior [72,73]. We therefore propose that iTBS neuromodulation may, in part, act by re-engaging this developmentally calibrated, experience-dependent gene program to promote synaptic recalibration. Future studies are needed to explore whether similar mechanisms are preserved or disrupted in neurodevelopmental conditions such as ASD with comorbid insomnia.

These findings are encouraging, but several important limitations should be acknowledged. First, the AHBA atlas has inherent constraints: only two donors provide right-hemisphere data, and all transcriptomic samples originate from neurotypical individuals rather than those with ASD. These factors may introduce bias when aligning transcriptional information with our neuroimaging results. Several avenues could strengthen future work. Second, although interest in overlapping system-level architecture is increasing, methodological approaches for characterizing this organization remain underdeveloped. The Shannon-entropy diversity coefficient applied here, for instance, requires further validation in independent datasets. Finally, while our multi-scale findings converge on a plausible mechanistic pathway, potentially involving modulation of receptor systems or components of the Ca²⁺–PKA axis during iTBS, direct experimental evidence will be necessary to definitively establish causality.

## Conclusions

This study establishes a multi-scale link among the Shannon-entropy diversity coefficient, iTBS-induced network modulation, and intrinsic molecular architecture in ASD with insomnia. Our findings suggest that iTBS may improve sleep in ASD through reorganization of overlapping system-level architecture in the brain, a process likely shaped by underlying chemoarchitectural constraints and involving Ca²⁺–PKA axis–related synaptic tuning genes. These insights offer a novel perspective on the neurobiological mechanisms of iTBS and may guide the development of biomarker-informed neuromodulation strategies tailored to individual neurochemical and genetic profiles.

## Supporting information

Glossary

navigation

## CRediT authorship contribution statement

**Huashuang Zhang :** Writing-original draft, Conceptualization, Investigation, Formal analysis, Data curation, Funding acquisition, Resources. **Xiong Bincan:** Data curation, Visualization, Software, Methodology, Investigation, Supported data analyses, Resources. **Hongchi Liu:** Writing-review & editing, Survey design, Software, Data quality assurance, Resources. **Jinwen Zhang:** Review & editing, Validation. **Ruimin Yu:** Review & editing, Resources, Validation. **Wenqi Jiang:** Writing-review & editing, Validation. **Jiongjia She:** Writing-review & editing, Resources. All authors read and approved the final submitted manuscript.

## Ethics approval and consent to participate

Ethics approval was granted by Foshan University (approval number FUME2024013) and Third People’s Hospital of Songzi (202404XA11). This study was conducted in accordance with the ethical principles of the Declaration of Helsinki. All participants provided written informed consent.

## Acknowledgment

We would like to thank all participants who took part in our study.

## Data availability

The datasets used and/or analyzed during the current study are available from corresponding author on reasonable request.

## Consent for publication

Not applicable.

## Glossary

ASD: autism spectrum disorder
iTBS: intermittent theta-burst stimulation
ISI: insomnia Severity Index
fMRI: functional magnetic resonance imaging
PPI: protein–protein interaction
LTP: long-term potentiation
E/I: excitation and inhibition
PET: positron emission tomography
RMT: resting motor threshold
OFC: orbitofrontal cortex
TR: repetition time
TE: echo time
FD: framewise displacement
CSF: cerebrospinal fluid
MNI: montreal Neurological Institute
FC: functional connectivity
MVPA: multivariate pattern analysis
SVR: support vector regression
LOOCV: leave-one-out cross-validation
AHBA: Allen Human Brain Atlas
log₂FC: log₂ fold-change
MDD: major depressive disorder
SCZ: schizophrenia
BD: bipolar disorder
IBD: inflammatory bowel disease
GO: Gene Ontology
TSEA: Tissue Specific Expression Analysis
CTEA: Cell Type-Specific Expression Analysis
*PPI*: Protein–protein interaction
Ctx.Glt25d2: cortical excitatory neurons
Cb.Pcp2: cerebellar Purkinje cells
BP: biological process
CC: cellular component
MF: molecular function
PKA: protein kinase A
3Di: Developmental, Dimensional and Diagnostic Interview
ABC: Antecedents, Behaviours and Consequences (chart)
ACE: ADHD Child Evaluation
CONT: control
DEFAULT: default mode
DORSATTN: dorsal attention
LIMBIC: limbic
SALVENTATTN: salience/ventral attention
SOMMOT: somatomotor
TEMPPAR: temporal–parietal
VISCENT: visual central
VISPERI: visual peripheral
MGluR5: glutamatergic
GABAA: GABAergic

