## Supplementary material for "Intermittent Theta Burst Stimulation Modulates Overlapping Network Architecture Through Neurochemical and Gene-Regulatory Pathways in Autism Spectrum Disorder With Insomnia": Figure captions

**Fig 1.** Schematic overview of the study design and analytical workflow.

**Fig 2.** Schematic representation of the functional brain networks. (A) Schematic_pre. (B) Schematic_post (Specific_Networks highlighted). The atlas comprises 200 regions of interest assigned to 17 functional networks. Each network is coded by a distinct color, and node size reflects its degree within the network.

**Fig 3.** Distributional alterations in the Shannon-entropy diversity coefficient across 13 overlapping brain regions following 8 weeks of neuronavigated iTBS (FDR-corrected, *p* < 0.05). Pre–post differences in the diversity coefficient are shown for (A) the sham iTBS and (B) the real iTBS.

**Fig 4**. Associations between altered Shannon-entropy diversity coefficients in ASD with insomnia and specific neurotransmitter architectures, including (A) DRD2; (B) DAT; (C) mGluR5; (D) VAChT; (E) 5-HT1A; (F) GABA_A._ DRD2, Dopamine D2 Receptor; DAT, dopamine transporter; mGluR5, metabotropic glutamate receptor 5; VAChT, vesicular acetylcholine transporter; 5-HT1A, 5-Hydroxytryptamine receptor 1A; GABA_A_, gamma-aminobutyric acid type A.

**Fig 5**. Differential expression patterns across disorders. Scatter plots illustrate the relationship between imaging–gene association weights and transcriptomic dysregulation for six conditions: (A) ASD; (B) MDD; (C) SCZ; (D) BD; (E) Alcohol dependence; (F) IBD. ASD, autism spectrum disorder; MDD, major depressive disorder; SCZ, schizophrenia; BD, bipolar disorder ; IBD, inflammatory bowel disease.

**Fig 6**. Gene-enrichment analyses for the differential overlapping brain regions. (A) Functional enrichment of related genes. The x-axis shows gene counts; the y-axis lists functional categories. Bubble area reflects the number of overlapping genes; bubble color denotes −log10(FDR). p values were adjusted using the FDR method. BP: biological process (*p* < 0.05, FDR-corrected); CC: cellular component (FDR corrected, *p* < 0.05); MF: molecular function (uncorrected, *p* < 0.05). (B) Tissue-specific expression, plotted as −log10(P). (C) Cell-type-specific expression, shown as −log10 (P). (D) Temporal expression trajectories.

**Fig 7**. PPI network constructed from genes associated with the differential overlapping brain regions. (A) The PPI network includes 64 genes with PRKACB, PRKACA, and PRKACG identified as the highest-degree hub genes. *p* values represent the significance of enrichment for proteins encoded by these genes. (B–D) Spatiotemporal expression profiles of PRKACB, PRKACA, and PRKACG. The x-axis depicts developmental time points; the y-axis reflects gene signal intensity. The lower panels show gene expression in neocortical regions at corresponding stages. PPI, Protein–protein interaction.
