## Supplementary material for "Intermittent Theta Burst Stimulation Modulates Overlapping Network Architecture Through Neurochemical and Gene-Regulatory Pathways in Autism Spectrum Disorder With Insomnia": Table S1

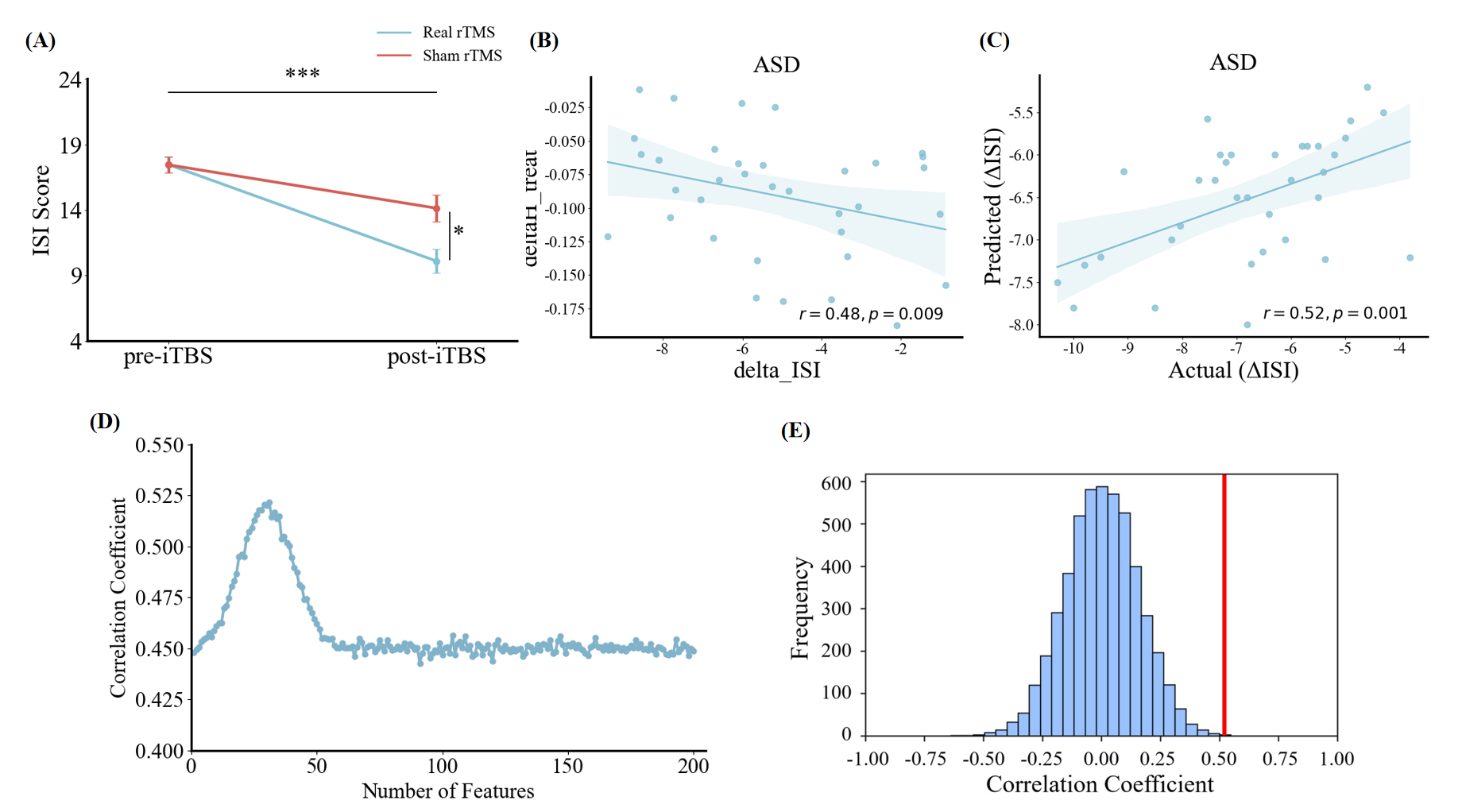

**Fig S1.** Clinical outcome measures and SVR prediction results. (A) Group-level changes in Insomnia Severity Index (ISI) scores before and after treatment, showing a significantly greater reduction in the real iTBS group compared with the sham group.
(B) Association between the iTBS-induced change in Shannon-entropy diversity within visual-pathway regions (ΔH_treat) and the observed improvement in ISI scores (ΔISI). (C) Scatter plot showing the relationship between SVR-predicted and actual ISI improvement, demonstrating significant predictive performance (r = 0.52). (D) Feature-weight distribution illustrating the contribution of entropy-based features within the SVR model. (E) Permutation-based null distribution (1,000 iterations) of correlation coefficients, with the red line marking the true model estimate, indicating that the prediction accuracy was unlikely due to chance (*p* = 0.001). Support vector regression, SVR.
