## Supplementary material for "Intermittent Theta Burst Stimulation Modulates Overlapping Network Architecture Through Neurochemical and Gene-Regulatory Pathways in Autism Spectrum Disorder With Insomnia": navigation

### TMS neuronavigation

To enhance anatomical precision and minimize interindividual variability, neuronavigation was implemented using the Navigator software, enabling stereotaxic registration of each participant’s brain with the TMS coil (Julkunen et al., 2009; Ruohonen et al., 2010). During baseline or sham sessions (conducted prior to MPRAGE acquisition), a standardized MNI template guided the navigation, whereas for the active iTBS session, each participant’s individual MPRAGE image was employed to ensure maximal anatomical accuracy.

The left OFC stimulation site was localized in three dimensions following well-established anatomical landmark protocols (Crespo-Facorro et al., 1999). Initially, the EEG International 10–20 system was used to identify position Fp1, a procedure that compensates for variations in skull size and is routinely applied in clinical TMS when neuronavigation is unavailable. The coil was first positioned tangentially to the scalp at Fp1, and the resulting focal point of stimulation was visualized on the participant’s structural MRI within the neuronavigation system. A digital marker was then placed at this focal point, and its coordinates were verified to ensure that they corresponded to the intended left OFC/frontopolar cortex (Brodmann area 10). If the Fp1-based position did not accurately align with the target region, the coil was gradually adjusted inferiorly and/or laterally until the projected stimulation site was located within Brodmann area 10.

The neuronavigation procedure aimed to deliver stimulation to BA10 as close as possible to the adjacent BA11. However, precise placement in more ventral regions was physically constrained by the geometry of the coil and individual head morphology (e.g., forehead curvature and nasal bridge distance). Consequently, the final scalp positions and MNI coordinates of the stimulation foci showed slight interindividual variation. Nevertheless, across participants, the focal point consistently localized just superior to the left eye, near or over the brow line. Sensitivity analyses confirmed that these small coordinate variations had negligible influence on neural or behavioral results (see Neuronavigational target coordinates section below). Figure S1 illustrates a representative electric field distribution induced by TBS over the left OFC/frontopolar cortex, simulated using SimNIBS 2 (Windhoff et al., 2013) with default head and coil models.

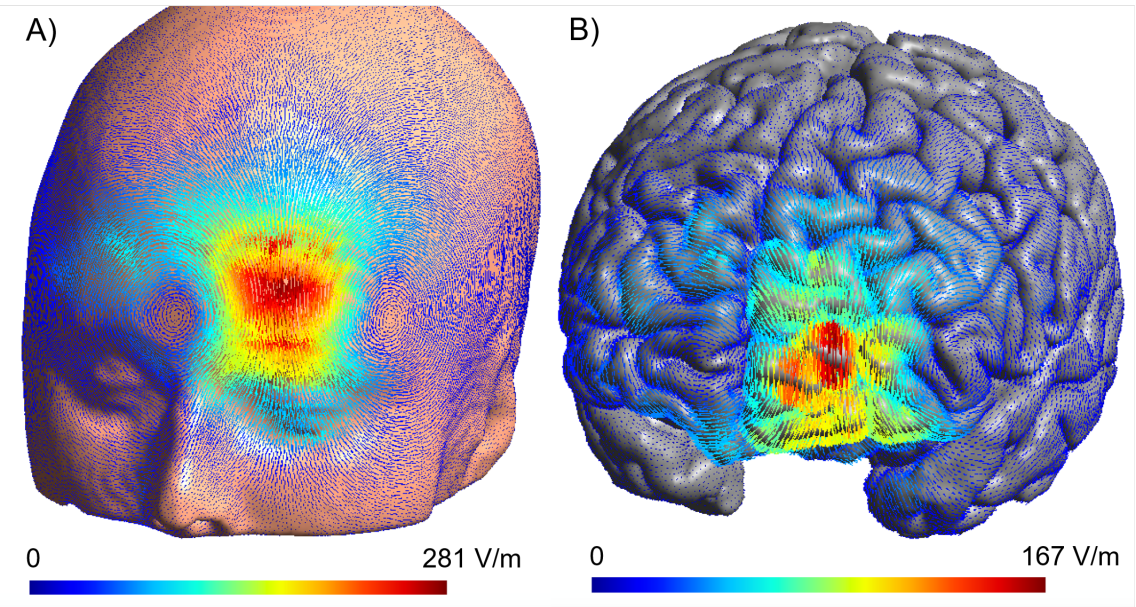

**Figure S1.** Computer simulation of the scalp distribution of the electric field induced by TBS over the left orbitofrontal/frontopolar cortex. The TMS coil placement approximated the targeting paradigm in the study. The TMS pulse amplitude was 100% of the maximum stimulator output. The simulation was carried out with SimNIBS 2 [<https://simnibs.github.io/simnibs/build/html/index.html>; (Windhoff et al., 2013)] using the default head model, tissue conductivity values, and MCF-B65 coil model.

**Neuronavigational target coordinates**

For each participant, the neuronavigation system provided the three-dimensional MNI coordinates of the stimulation focal point during the active iTBS session. To evaluate whether individual differences in stimulation loci contributed to outcome variability, we correlated the x-, y-, and z-coordinates with post-treatment changes in the Shannon-entropy diversity coefficient (ΔH) and ISI scores (ΔISI), analyzed separately for the iTBS and sham groups.

No systematic or spatially consistent associations were observed between target coordinates and either neural or behavioral outcomes. Coordinate variability in any axis (x, y, z) did not significantly relate to post-treatment H values or their change from baseline, nor to ΔISI scores in either group. These findings indicate that **minor deviations in the neuronavigational focal point had negligible influence on both network-level entropy measures and behavioral performance**, confirming the spatial robustness of the left OFC targeting. Moreover, the high reproducibility of results across participants supports the reliability of the iTBS stimulation protocol and suggests that observed treatment effects primarily reflect neurophysiological modulation rather than geometric variability in coil placement.
