## Supplementary material for "Intermittent Theta Burst Stimulation Modulates Overlapping Network Architecture Through Neurochemical and Gene-Regulatory Pathways in Autism Spectrum Disorder With Insomnia": Table S1

64 genes whose expression levels were significantly correlated with the spatial pattern of iTBS-induced alterations in the overlapping brain system (FDR corrected, *p* < 0.05)

AKAP5

CACNA1C

CACNA1D

CACNA1F

CACNA1S

CACNA2D1

CACNA2D4

CACNB3

CACNB4

CACNG2

CACNG3

CACNG4

CACNG6

CACNG7

CACNG8

CALM1

CALM2

CALM3

CAMK2A

CAMK2B

CNIH2

CNIH3

DGKZ

DLG1

DLG3

DLG4

DLGAP1

GRIA1

GRIA2

GRIA3

GRIA4

GRID1

GRIN1

GRIN2A

GRIN2B

GRIN2C

GRIN2D

GRIN3A

GRIN3B

HSPA2

INPP5A

IPMK

ITPKA

KIF13B

KIF5A

KIF5B

KIF5C

LYN

MINPP1

NOS1

PIKFYVE

PIP4K2A

PLCB4

PPFIA2

PPP1CA

PPP1CB

PPP1CC

PRKACA

PRKACB

PRKACG

RASGRP3

SHANK1

SHANK2

SHANK3
