## Supplementary material for "Intermittent Theta Burst Stimulation Modulates Overlapping Network Architecture Through Neurochemical and Gene-Regulatory Pathways in Autism Spectrum Disorder With Insomnia": Table 1

| **Table 1** Demographics and clinical characteristics. | | | | |
| --- | --- | --- | --- | --- |
| Characteristics | Real stimulation  (n = 35) | Sham stimulation  (n = 35) | t/*x*^2^ | *p* |
| Age (years) | 33.78 ± 11.22 | 34.14 ± 11.25 | -0.13 | 0.890 |
| Gender |  |  | 1.00 | 0.317 |
| Male | 20 | 25 |  |  |
| Female | 15 | 10 |  |  |
| FSIQ | 116.93 ± 9.12 | 120.69 ± 14.23 | -1.32 | 0.190 |
| ADOS | 10.80 ± 3.91 | 11.51 ± 4.13 | 0.74 | 0.460 |
| Insomnia duration (years) | 9.25 ± 10.72 | 10.21 ± 11.20 | -0.37 | 0.720 |
| Dairy quantity of hypnotics used (diazepam equivalent, mg) | 1.30 ± 2.48 | 1.56 ± 2.82 | -0.41 | 0.680 |
| FSIQ, full-scale intelligence quotient; ADOS, Autism Diagnostic Observation Schedule. | | | | |
